# Use of whole genome sequencing to identify low-frequency mutations in COVID-19 patients treated with remdesivir

**DOI:** 10.1101/2022.11.20.22282552

**Authors:** Kuganya Nirmalarajah, Finlay Maguire, Winfield Yim, Patryk Aftanas, Angel X. Li, Altynay Shigayeva, Lily Yip, Xi Zoe Zhong, Allison J. McGeer, Samira Muberka, Robert Kozak

## Abstract

**Background:** We investigate the effects of remdesivir (RDV) treatment on intra-host SARS-CoV-2 diversity and low-frequency mutations in moderately ill hospitalized COVID-19 patients and compare them to patients without RDV treatment.

**Methods:** Sequential collections of nasopharyngeal and mid-turbinate swabs were obtained from 16 patients with and 31 patients without RDV treatment. A total of 113 samples were sequenced and mutation analyses were performed.

**Results:** We did not identify any drug resistant mutations during RDV therapy. In genes encoding and associated with the replication complex, low-frequency minority variants that do not reach fixation within the sampling period were detected in 6/16 (37.5%) and 14/31 (45%) patients with and without RDV treatment respectively. We did not detect significant differences in within-host diversity and positive selection between the RDV-treated and untreated groups.

**Conclusions:** Minimal intra-host variability and stochastic low-frequency variants detected in moderately ill patients suggests little selective pressure in patients receiving short courses of RDV. Patients undergoing short regimens of RDV therapy should continue to be monitored.

## Background

The global pandemic from SARS-CoV-2 has resulted in millions of deaths, and hundreds of millions of cases. While vaccines have been shown to have high effectiveness against the variants of concern in preventing severe disease [1], there have been breakthrough infections, and some populations remain at risk for severe disease.

SARS-CoV-2 specific small molecule inhibitors offer a promising means of reducing hospitalizations and mortality. Remdesivir (RDV) is a nucleotide analogue that is administered by the intravenous route as a prodrug, and subsequently converted by the host cell into the active triphosphate form to inhibit viral replication [2].

RDV has been an important therapeutic option against SARS-CoV-2 and has maintained activity against all variants that have emerged so far [3]. Therapeutic efficacy demonstrated by increased survival has been noted in hospitalized patients following 5 days of treatment, and 3 days in out-patient trials [4, 5]. Known resistance conferring mutations to RDV are very rare based on a survey of SARS-CoV-2 genomes available in GISAID [6]. To date, RDV resistance has been primarily reported in immunocompromised patients who are on prolonged therapy [7, 8] Additionally, a recent report highlighted an immunocompromised patient who had rebound infections and multiple courses of RDV, but no resistance mutations were identified [9]. Overall, there is a paucity of data on patients with serial sample collections over the course of their treatment making it difficult to determine when, or if, resistance mutations arise, and at what frequency.

Hight-throughput sequencing is a technique that has the capacity to identify low-frequency minority variants that would not be identified by conventional Sanger sequencing. This has been shown to be useful for detecting low-frequency mutations that emerge following the administration of antiviral therapy for numerous viruses [10, 11]. Inadequate therapy can be a driver for the selection of drug resistance, and current recommendations are for 10 days of RDV therapy for hospitalized patients. While mutations conferring resistance to RDV have been identified in patients who received prolonged therapies for SARS-CoV-2 [7, 12], it remains unclear if hospitalized individuals who received shorter courses of therapy are likely to develop mutations that confer resistance to RDV.

In this study we collected serial samples from patients who had short a RDV therapy duration. Novel polymorphisms identified as low-frequency variants were analyzed for their potential resistance profiles and tested for selection.

## Methods

### Patients and sample collection

Serial samples obtained from hospitalized adult patients with laboratory-confirmed COVID-19 were recruited from 5 different hospital sites in the greater Toronto area. Patients were prospectively recruited by the Toronto Invasive Bacterial Disease Network (TIBDN) from March 2020 to April 2022 for a series of studies investigating viral shedding and environmental contamination over time. Informed consent was obtained and all participating TIBDN hospitals were granted research ethics approval by their respective ethics boards (REB #2218). Demographic and clinical data were obtained from electronic medical record review and patient interview. Serial nasopharyngeal or mid-turbinate samples were collected approximately every 3 days starting from date of study enrollment until recovery, refusal, or death.

### Whole genome sequencing

The ARTIC SARS-CoV-2 protocol (https://artic.network/ncov-2019) was used for amplicon generation. Sequencing was performed using the methods outlined [13, 14]. Briefly, nucleic acid extraction was performed using the EasyMag platform (Biomeurieux) [15]. Next, cDNA was generated according to the ARTIC protocol. After cDNA synthesis, two multiplex PCR tiling reactions were prepared using the ARTIC V3, V4, and V4.1 primers (depending on sampling date). In a PCR tube, cDNA was combined with Q5 High-Fidelity 2X Master Mix (NEB, USA). To mix #1, 10 µM of ARTIC primer pool #1 were added and 10 µM of ARTIC primer pool #2 were added to mix #2. PCR cycling was then performed as follows: 98 °C for 30 s followed by 35 cycles of 98 °C for 15 s and 65 °C for 5 min. Following this reaction, PCR products were combined, and a clean-up with AMPure XP beads was performed (Beckman Coulter, USA). The quantity of amplicons was measured with the Qubit 4.0 fluorometer using the 1X dsDNA HS Assay Kit (Thermo Fisher Scientific, USA). The sequencing libraries were prepared using the Nextera DNA Flex Prep kit (Illumina, USA) as per manufacturer’s instructions. Paired-end (2×150bp) sequencing was performed on a MiniSeq with a 300–cycle reagent kit (Illumina, USA).

### Bioinformatic analysis

Short-read sequence processing, genome assembly, and Pangolin lineage assignment were done using the SIGNAL v1.5.6(SARS-CoV-2 Illumina GeNome Assembly Line) pipeline (https://github.com/jaleezyy/covid-19-signal) [14]. Within SIGNAL, the consensus genome sequences are generated based on the SARS-CoV-2 reference genome (MN908947.3) using BWA-MEM [16] v0.7.17. PANGO [17] lineages were assigned using pangolin [18] v3.1.17 with PangoLEARN 2022-01-20 and coverage statistics were calculated with BEDTools [19] v2.26.0. Annotated amino acid mutations were predicted by Breseq v035.6 using default parameters including cut-offs of 0.05 for minimum polymorphism frequency and 2 for polymorphism minimum coverage [20]. Breseq results were visualized using the pandas [21] v1.5.0, matplotlib [22] v3.5.3, and seaborn [23] v0.11.2 packages via Python v3.10.7.

### Phylogenetic analysis

Transition (Ti) and transversion (Tv) substitutions, deletions, and insertions were tabulated from Breseq results. Hypothesis Testing using Phylogenies (HyPhy) v2.5.42 was used for testing signals of positive selection [24] in the ORF1ab gene on RDV-treated sequences. Prior to selection analysis, Virulign [25] v1.0.1 was used to generate codon alignments of all sequences with the SARS-CoV-2 (MN908947.3) ORF1ab as the reference. A HyPhy inferred tree was generated and phylogenies were labelled using the Phylotree Widget. Excess stop codons were removed using HyPhy before performing selection analysis. The adaptive branch-site random effects likelihood (aBSREL) [26] method from HyPhy was used to test for evidence of positive selection amongst the foreground RDV branches relative to the untreated sequences.

### Statistical analysis

Statistical testing and plotting were performed using RStudio v1.4.1717 with the dplyr [27] v1.0.10 and ggplot2 [28] v3.3.6 packages. The Mann Whitney U test was used to compare the distribution of mutation counts, and transition and transversion mutation types between patients with and without RDV treatment. The Chi-Square test was used to assess the difference between the Ti/Tv ratios in the RDV-treated and untreated groups. Statistical significance was determined based on a *P* value of less than 0.05.

## Results

### Cohort description

In total, 47 patients who were hospitalized from March 2020 to April 2022 and had at least two SARS-CoV-2 positive nasal samples (diagnostic or serial collections) were included in the study. Patient demographics and clinical characteristics are shown in Table 1. The median age of patients was 74 years (interquartile range [IQR], 61.5 - 84.25) and the median duration of hospitalization was 14 days (IQR, 10.25 - 22.75). Sixteen (34.8%) patients received at least one dose of RDV for a median duration of 4 days (IQR, 8.75 - 15). Each patient was sampled a median of 2 (IQR, 2-3) times following commencement of therapy and collection times are shown in Table 2. Serial collections from all patients were sequenced if SARS-CoV-2 RT-PCR cycle threshold values for the E-gene target were < 30. A total of 113 complete genomes were generated from 47 patients. Patients were infected with early B.1, B.1.1, and B.1.1.181 SARS-CoV-2 lineages and variants of concern (VOCs) including Alpha, Delta, and Omicron. No patients received additional antivirals in hospital following RDV treatment.

**Table 1:** Patient demographics and clinical characteristics of cohort.

|  | **Total (n = 47)** | **Remdesivir (n = 16)** | **No Remdesivir (n = 31)** |
| --- | --- | --- | --- |
| Median age, years (IQR) | 74 (61.5 - 84.25) | 71 (56.25 - 82) | 75 (63 - 89) |
| ≥ 65 years | 29 (61.7%) | 8 (50%) | 21 (67.7%) |
| ≤ 64 years | 18 (38.3%) | 8 (50%) | 10 (32.3%) |
| Male sex | 29 (61.7%) | 8 (50%) | 21 (67.7%) |
| Median duration of hospitalization (IQR) | 14 (10.25-22.75) | 11 (8.75 - 15) | 17 (12-33.5) |
| Median duration of RDV treatment (IQR) |  | 4 (2 - 9) |  |
| ICU admission | 2 (4.3%) | 2 (12.5%) | 0 |
| Mortality | 7 (14.9%) | 0 | 7 (22.6%) |
| Obesity | 2 (4.3%) | 1 (6.3%) | 1 (3.2%) |
| Diabetes | 16 (34%) | 7 (43.8%) | 9 (29%) |
| Cardiac disease | 18 (38.3%) | 6 (37.5%) | 12 (38.7%) |
| Vascular disease | 27 (57.4%) | 9 (56.3%) | 18 (58.1%) |
| Pulmonary disease | 17 (36.2%) | 6 (37.5%) | 11 (35.5%) |
| Renal disease | 9 (19.1%) | 4 (25%) | 5 (16.1%) |
| Cancer | 8 (17%) | 3 (18.8%) | 5 (16.1%) |
| Immunosuppressed | 5 (10.6%) | 3 (18.8%) | 2 (6.5%) |

Abbreviations: IQR, interquartile range; ICU, intensive care unit

**Table 2:**
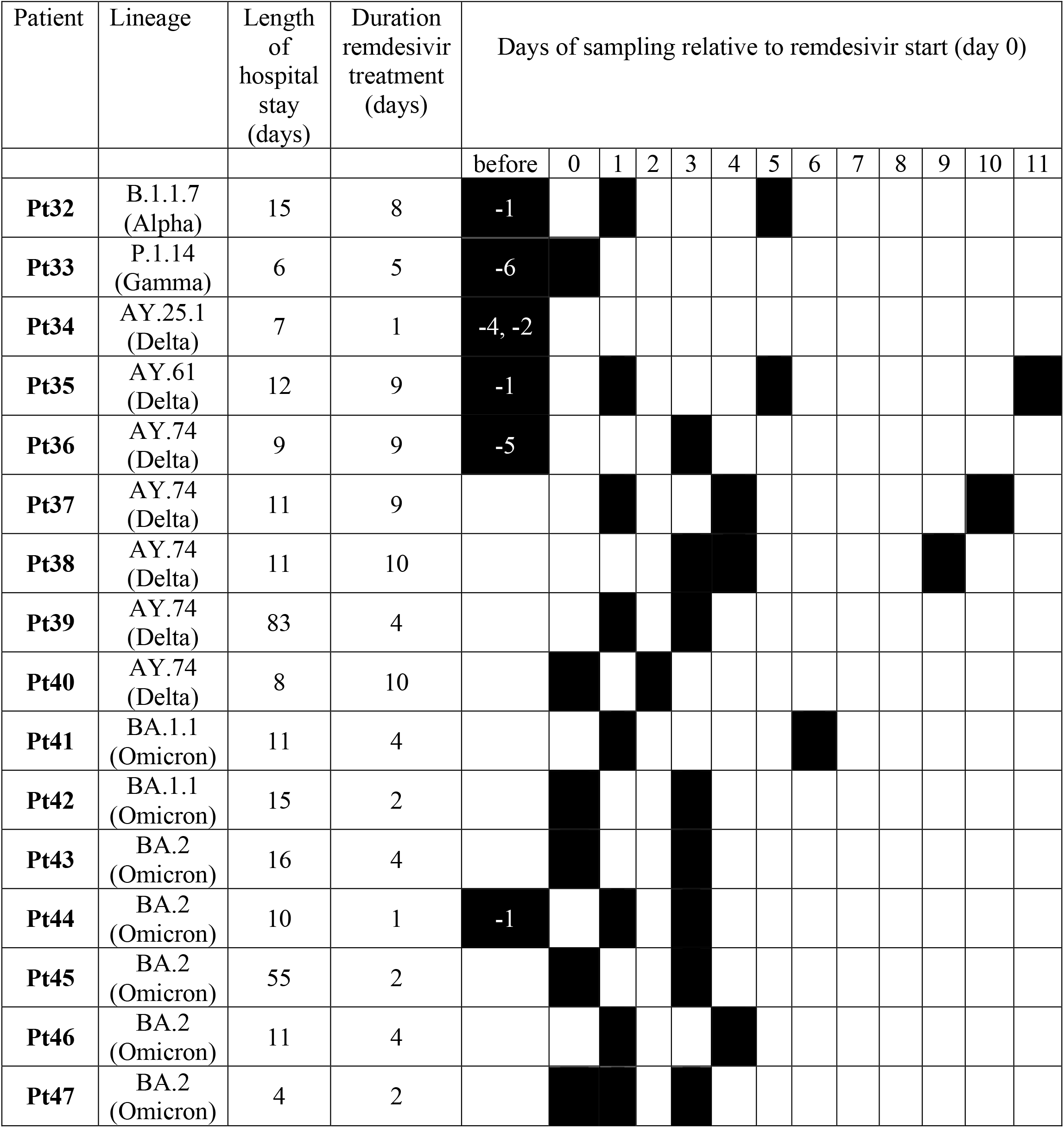
Timepoints collected for patients treated with RDV.

### Drug resistance mutations were not identified by WGS in RDV-treated patient cohort

Multiple mutations have been previously identified that confer varying levels of resistance to RDV. These drug-resistance mutations have been identified in genes associated with the viral replication complex (nsp7-nsp16) [6]. We examined 38 serial collections obtained from 16 RDV-treated patients. The consensus genome mutations from Breseq variant calling results from each timepoint for each patient were examined for drug-resistance mutations and none were detected. Notably nsp12 E802D, a drug-resistance mutations associated with treatment failure [7], was not detected as a consensus mutation in any patient samples regardless of the duration of RDV therapy. In parallel we analyzed 75 serial samples collected from 31 patients who did not receive RDV therapy and did not identify any drug-resistance mutations.

### Low-frequency variants identified during treatment

The highly sensitive sequencing platform allows for detection and quantification of low-frequency variants that would not be identified in consensus sequence. In vitro work done by other groups suggests that only mutations present at >15% were likely to represent viral adaptation in the face of selective drug pressure, thus we used a similar cut off [29]. Apart from majority variants present in consensus sequences, low-frequency minority variants within the replication complex and surrounding genes (nsp6 - nsp14) were identified in 6/16 (37.5%) of RDV-treated patients (Figure 1a). These low-frequency variants were unique and not repeatedly detected across patients infected with different lineages. Minority variants were identified in 14/31 (45%) of patients without RDV treatment (Figure 1b). Low-frequency variants identified in early time points did not persist in subsequent collections and did not become fixed in either group. Additionally, no low-frequency variants were observed at E802 or V166, both positions associated with RDV resistance in patients^7,12^. Low-frequency variants were also observed in other regions of the genome in both treatment groups (Supplementary Figure 1). No low-frequency variants were detected in the RNA dependent RNA polymerase (RdRp) palm domain.

**Figure 1.**
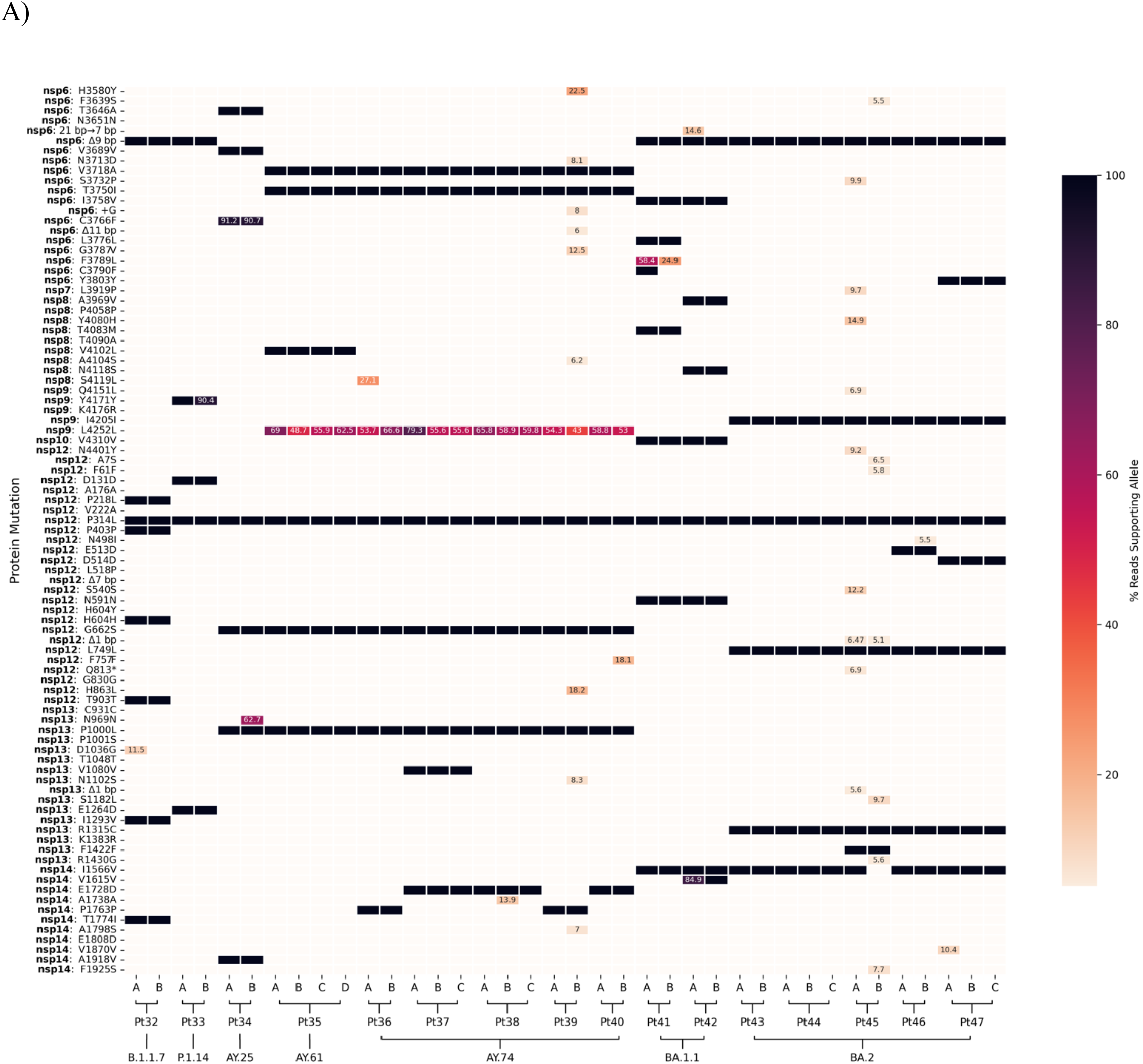
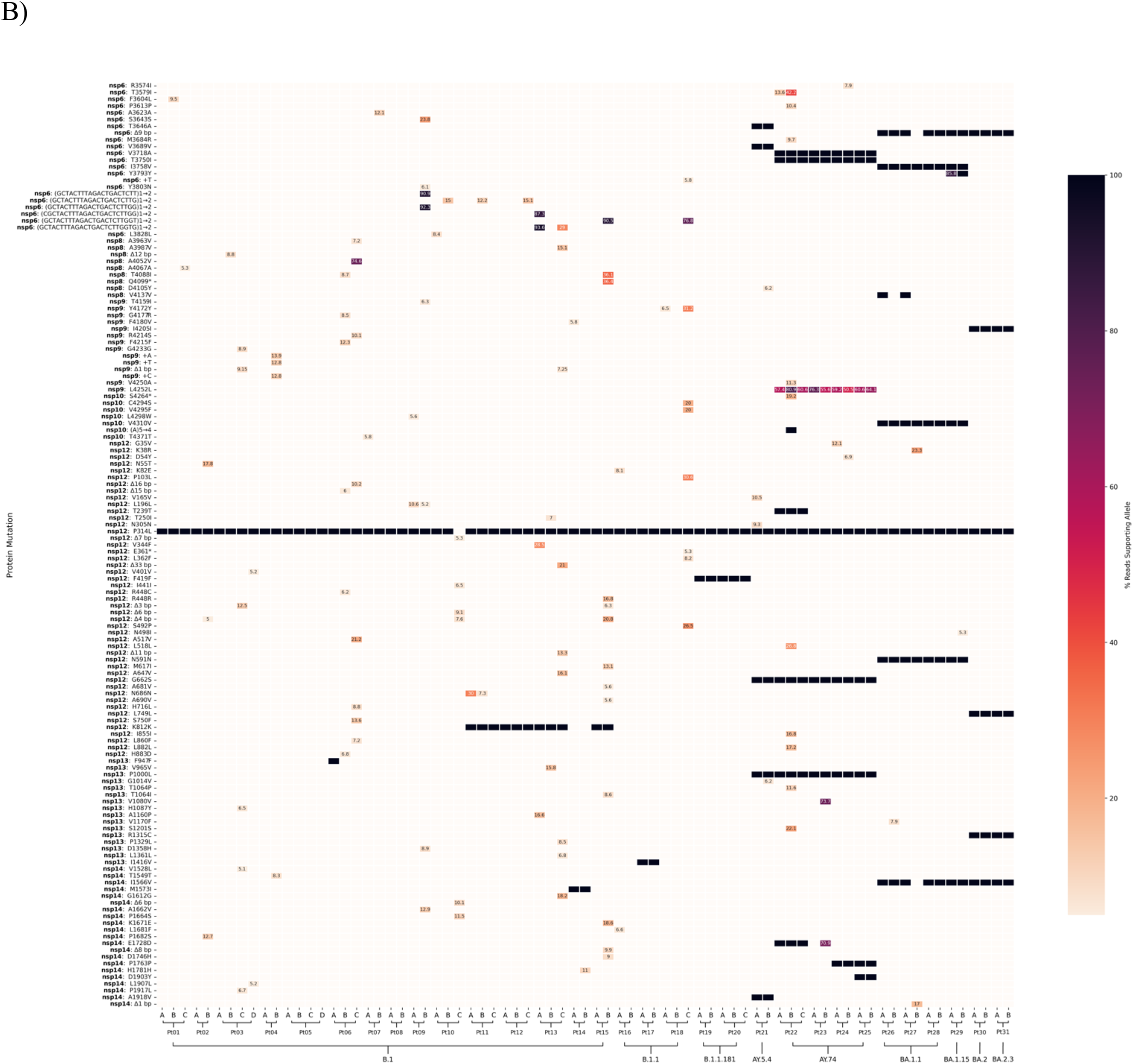
Mutation profile across nsp6-nsp14 for 39 serial swabs collected from RDV-treated patients (A) and 75 collected from untreated patients (B). Alphabetical ordering of patient serial samples represent collection date order. Mutations were analyzed and called against the reference sequence with the Breseq pipeline. Low-frequency minority mutations were retained and percentages are shown. Default parameters of Breseq including a 5% polymorphism frequency filter and a polymorphism minimum coverage of 2 were used.

### Within-host variability detected in RDV-treated and untreated patient cohorts

Viral genomic variability found within individuals infected with SARS-CoV-2 can result from the presence of quasispecies populations, co-infection, RNA degradation, or sequencing errors. We investigated intra-host viral diversity for patients treated with and without RDV therapy. Changes in the total number of amino acid mutations identified by Breseq, including synonymous and nonsynonymous single nucleotide polymorphisms (SNPs), insertions, deletions, and complex substitutions across samples from different time points can reveal intra-host viral diversity. Intra-host variability was detected in nsp6-nsp14 across serial collections in 10/16 (62.5%) patients treated with RDV (Figure 1a). However, there was no change in the consensus sequences of serial collections in 6/16 (37.5%) patients, indicating no intra-host diversity. Within-host diversity in nsp6-nsp14 was detected in 24/37 (64.68%) patients without RDV therapy (Figure 1b). There were 15/16 (93.75%) and 34/37 (91.89%) patients in the RDV-treated (Supplementary Figure 1a) and untreated (Supplementary Figure 1b) groups respectively where intra-host variation across serial collections were detected throughout the whole genome. Total within-host diversity was also compared amongst the RDV-treated and untreated groups by investigating the absolute difference in the number of total mutations per sample relative to the first analyzed serial collection (Figure 2). Within-host diversity between the RDV-treated and untreated groups in the nsp6 −14 region or the whole genome were not significantly different based on the Mann Whitney U test (Figure 3).

**Figure 2.**
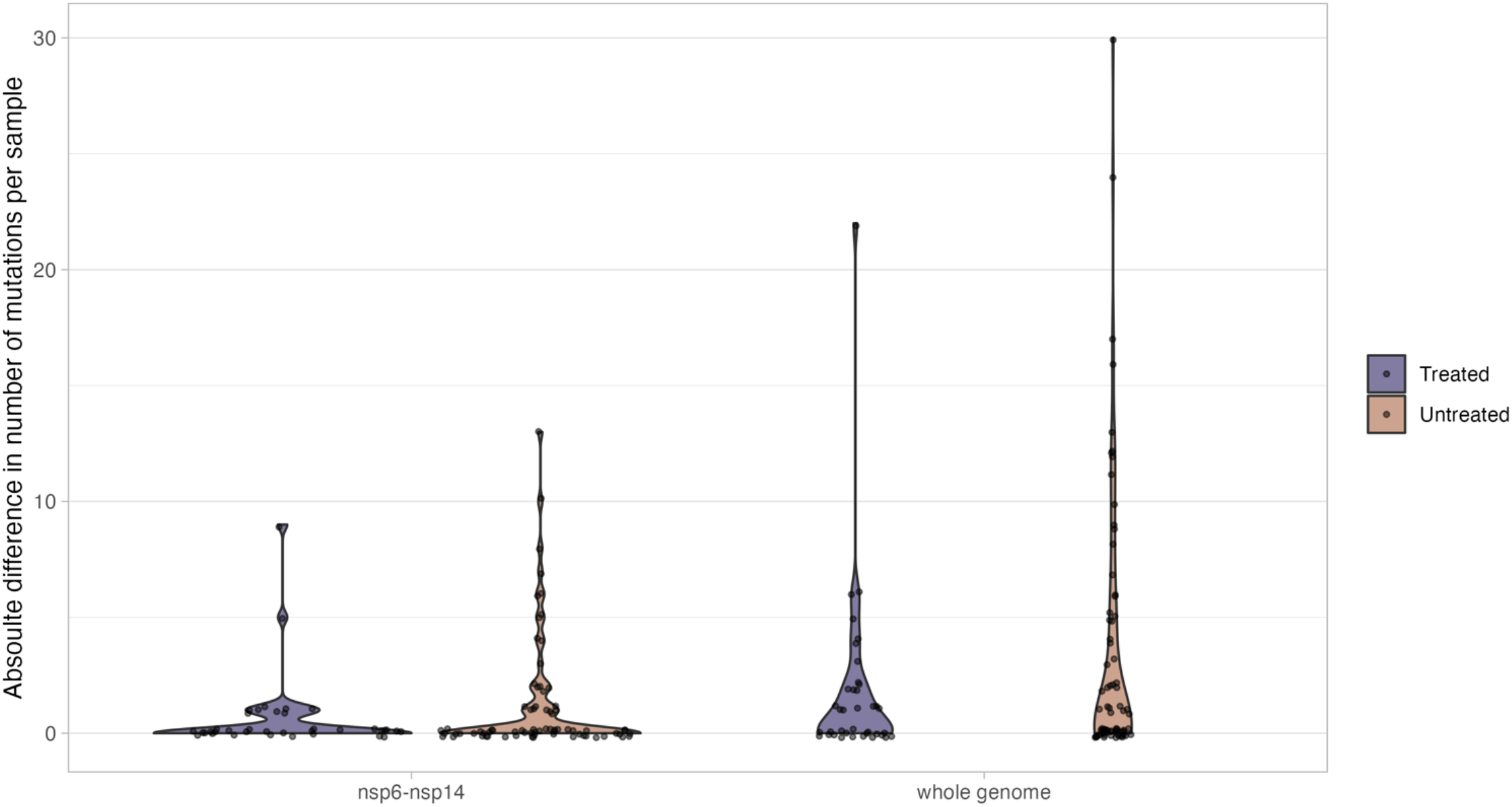
Absolute difference in number of total consensus and low-frequency mutations (including SNPs, amino acid insertions, amino acid deletions, and complex mutations) relative to the first examined sample of each patient within nsp6-nsp14 and the whole genome. The Mann Whitney U test was used to calculate *P* values comparing distributions between RDV-treated and untreated groups.

**Figure 3.**
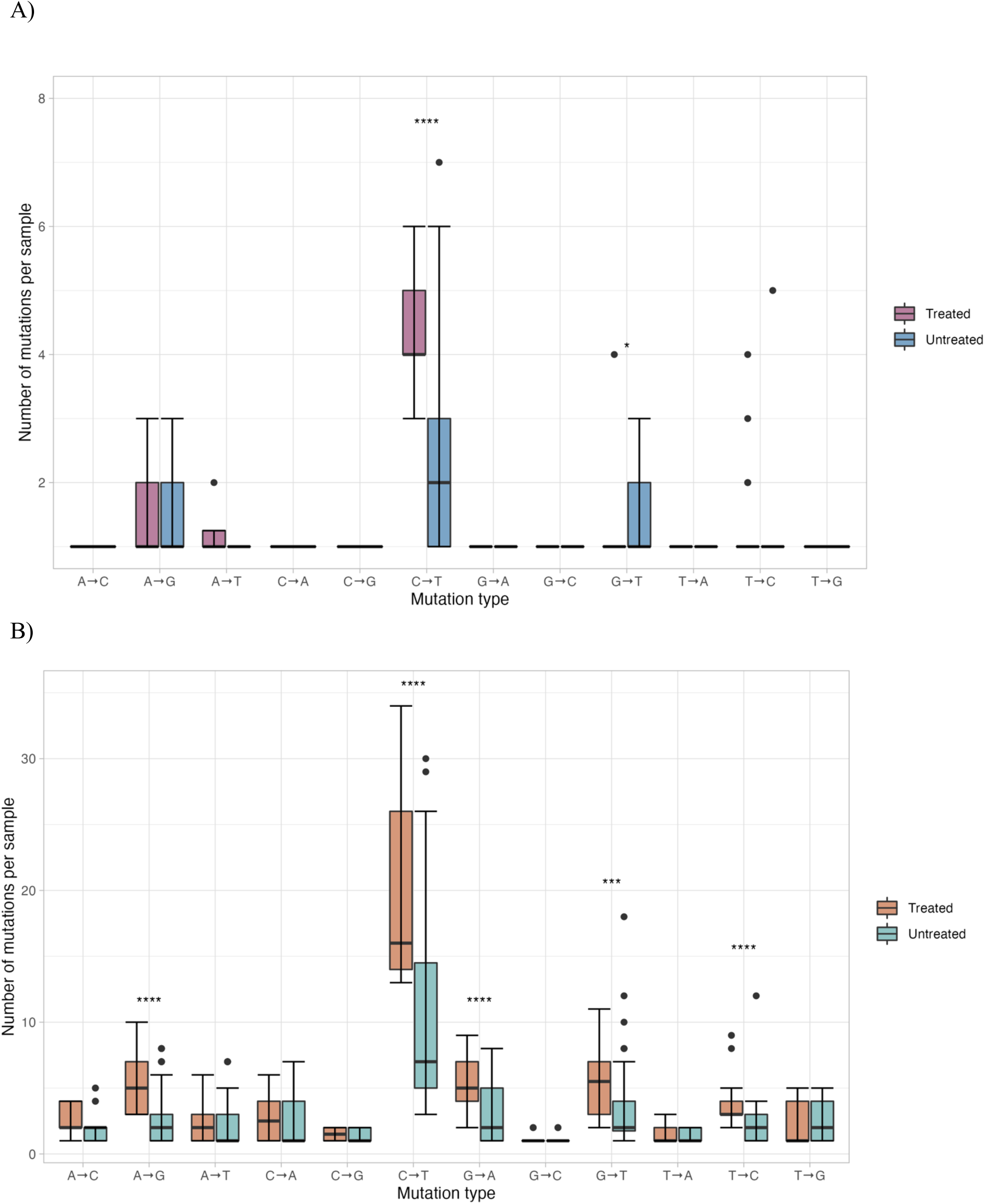
Boxplot summary of types of mutations representing transitions and transversions events in the RDV-treated and untreated groups within nsp6-nsp14 (A) and the whole genome (B). Points represent individual collections with counts of each substitution type for 38 samples from RDV-treated patients and 75 samples from patients without RDV treatment. The Mann Whitney U test was used to calculate *P* values comparing each mutation distribution between RDV-treated and untreated groups.

### Within-host variability and selection processes are minimal in RDV-treated and untreated patients

Transition (Ti) and transversion (Tv) events were examined across the whole genome for patients treated with and without RDV therapy. Transitions are substitutions interchanging bases of the same ring structure while transversions are interchanges of purine bases for pyrimidine bases. The total number of all transition and transversion events from each sample from all patients amongst the RDV-treated and untreated group were compared for the nsp6-nsp14 region (Figure 3a) and the whole genome (Figure 3b). C→T transitions were the most common type of substitutions in both RDV-treated and untreated groups. Prior to correcting for phylogenies, significant differences in the number C→T and G→T mutations in the nsp6-14 region (Figure 3a) and A→G, C→T, G→A, G→T, and T→C (Figure 3b) mutations in the whole genome between the RDV-treated and untreated group were observed. However, these differences can be attributed to the varying makeup of SARS-CoV-2 lineages in each treatment group, as the untreated group contains mostly early lineages. In the RDV-treated group, 1256 transitions and 605 transversions were identified within the whole genome, giving a Ti/Tv ratio of 2.09. There were 1258 transitions and 535 transversions in the untreated group within the whole genome with a Ti/Tv ratio of 2.35. When comparing the Ti/Tv ratios among the RDV-treated and untreated groups with the Chi-Square test, no significant difference was observed. Furthermore, we identified 164 small and mid-size deletions across the whole genome in the 16 RDV-treated patients of which 21 were low frequency (Supplementary Figure 1a). 190 deletions were detected in the 37 patients without RDV treatment with 59 low-frequency deletions (Supplementary Figure 1b). Deletions are denoted on heatmap figures by “Δ bp coding”. Most deletions detected were found in the consensus sequences of samples and are characteristic of VOCs particularly the Delta and Omicron lineages.

To further investigate whether selective pressures were exerted in the RDV-treated group, we tested for episodic positive selection while correcting for the phylogenetic structure of our samples. Codon alignments for ORF1ab were generated for all sequences and the adaptive Branch-Site Random Effects Likelihood (aBSREL) model [26] from HyPhy [24] was applied to an RDV-treatment labelled phylogeny generated by the Phylotree Widget. No evidence of episodic diversifying positive selection was identified in the ORF1ab gene in the foreground RDV branches relative to the untreated branches. Together, this suggests that evolutionary processes such as positive selection, replicase complex adaptation, and resistance emergence are minimal with RDV treatment.

## Discussion

In this cohort study, whole genome sequencing was used to identify low-frequency mutations in patients with serial collections during RDV therapy. Low-frequency variants were not detected at in nsp12 at E802, D484, V166 or at any of the other drug-resistance mutations positions reported previously [6-9]. Our findings suggest that nsp12 P314L is not associated with resistance to RDV in our patient population, as has been shown by others [9]. Overall, RDV did not result in the generation of low-frequency drug-resistance mutations in the short term. Martinot et al. (2021) identified the nsp12 D484Y mutation within the RdRp finger subdomain after RDV treatment and not before. Throughout longitudinal sampling, we did not observe any significant positive selection or any minority variants in genes encoding the replication complex (or throughout the genome) increase in frequency to apparent fixation. This suggests little selective pressure and minimal changes to viral intra-host variability as a result of RDV treatment. In RDV-treated patients, within nsp6-nsp14, we did not detect the persistence of low-frequency variants across timepoints from the same individuals or recurring variants at the same loci amongst different RDV patients. Additionally, within nsp6-nsp14, accumulation of *de novo* low, intermediate, or fixed variants throughout the course of infection in either the RDV-treated or untreated groups was not observed. Together, this suggests that low and intermediate frequency variants detected in samples from RDV patients arose sporadically and are likely random mutations rather than drug-resistance mutations. In contrast to our findings, Gandhi et al. (2022) reported the emergence and temporal increase in allele frequency of the *de novo* nsp12 E802D mutation in an RDV-treated immunocompromised patient after day 7 of therapy. To date, there have only been case reports of antiviral resistance being identified, and no one has performed serial sampling in a larger set of patients on RDV therapy with a comparison group.

Similar to Heyer et al (2022) we saw very little increase in variant allele frequencies and minimal intra-host diversity in the samples from RDV-treated patients. However, the authors reported increased genomic variability and 9 novel nucleotide variants (including 2 in the RdRp) that reached apparent fixation in a single patient with follicular lymphoma suffering from severe and prolonged infection and was treated with 2 courses of RDV [12]. Such observations suggest that RDV treatment can lead to increased intra-host genomic diversity and emergence of fixed mutations in immunodeficient hosts with prolonged infections. Furthermore, Heyer et al. argue that the emergence of novel variants cannot be attributed to prolonged infection alone as similar patients given other treatments such as anti-inflammatories resulted in sporadic, transient, and lower frequency variants. Existing studies suggest that within-host dynamics arise stochastically and that mutations that are advantageous to the virus may exist at low-frequency and emerge under selective pressure [12, 30, 31]. Our data suggest that if this is the case, strong selection is not present in the early phase of treatment. In our RDV-treated cohort, the majority did not have prolonged infections, mostly experienced moderate disease, and only 3/16 patients were immunocompromised. We speculate that minimal intra-host diversity and the emergence of low-frequency variants as observed in both our RDV-treated and untreated groups arise stochastically due to genetic drift and not from RDV selective pressures. Other studies in persistently infected immunocompromised patients have also failed to identify resistance mutations to RDV [32], although interestingly, mutations that reduce monoclonal antibody neutralization efficacy arise more easily [33, 34].

Our study was different from previously published studies as it focused on patients who received less than the recommended 10 days of RDV therapy and indicates that this does not select for resistant viruses in hospitalized patients. This is indirectly supported by the fact that RDV has been used as 3-day treatment for non-hospitalized patients, and this has not resulted in documented resistance emerging [5]. While it cannot be discounted that other mutations arose at later time points, this seems unlikely as all RDV-treated patients recovered and were discharged, none having prolonged infection. The findings in our study provide additional insight into the impact of RDV treatment on intra-host genomic variation and the emergence of novel variants. We highlight these effects in moderately ill patients with shorter-term infection and without severe immunocompromise. Although 5-day courses of RDV for moderately ill hospitalized patients [35], 3-day regimens for at risk non-hospitalized patients, and the use of other antivirals such as nirmatrelvir+ritonavir and molnupiravir have been shown to be effective, it is important to continue to monitor for genomic variability in patients with both short-term and prolonged infections. Considering that resistance has not been reported following 3- or 5-day course of RDV, it suggests that the barrier to RDV resistance is high. It is likely that immunosuppression of the host is needed for the development of resistance.

Our study had several limitations. These included the small size of the cohort, and that only patient samples with sufficiently high viral load (e.g. cycle threshold value <30) were suitable for sequencing. As a result, our sampling periods, number of samples per patient, and sampling intervals varied throughout the study. The phylogenetic makeup of our treatment groups also varied. RDV-treated patients were infected with VOCs mainly Delta and Omicron, whereas most untreated patients experienced infection with early SARS-CoV-2 lineages. We did attempt to control for the phylogenetic structure while testing for positive selection, however, this will still reduce our statistical power. The difference in the number of patients in each treatment is another limitation of our study. While our treatment groups have comparable demographic and clinical characteristics, we could not account for variability in immune pressures within individual hosts. Furthermore, although short read sequencing is a robust tool, it is not completely free from error or false positivity. Artefactual low-frequency minority variants within the treated and untreated patient groups have not been ruled out. Future improvements to whole-genome sequencing assays are needed to better interrogate samples with low viral loads and improve detection of antiviral resistance mutations.

## Data Availability

All data produced in the present study are available upon reasonable request to the authors

## Acknowledgments

We thank all COVID-19 patients who consented to participate in this research. Thank you to the Toronto Invasive Bacterial Disease Network for all laboratory, recruitment, and research ethics contributions. We acknowledge the Shared Hospital Laboratory for their SARS-CoV-2 genome sequencing facilities and support.

