## Supplementary Figure 1 for "Use of whole genome sequencing to identify low-frequency mutations in COVID-19 patients treated with remdesivir"

A)

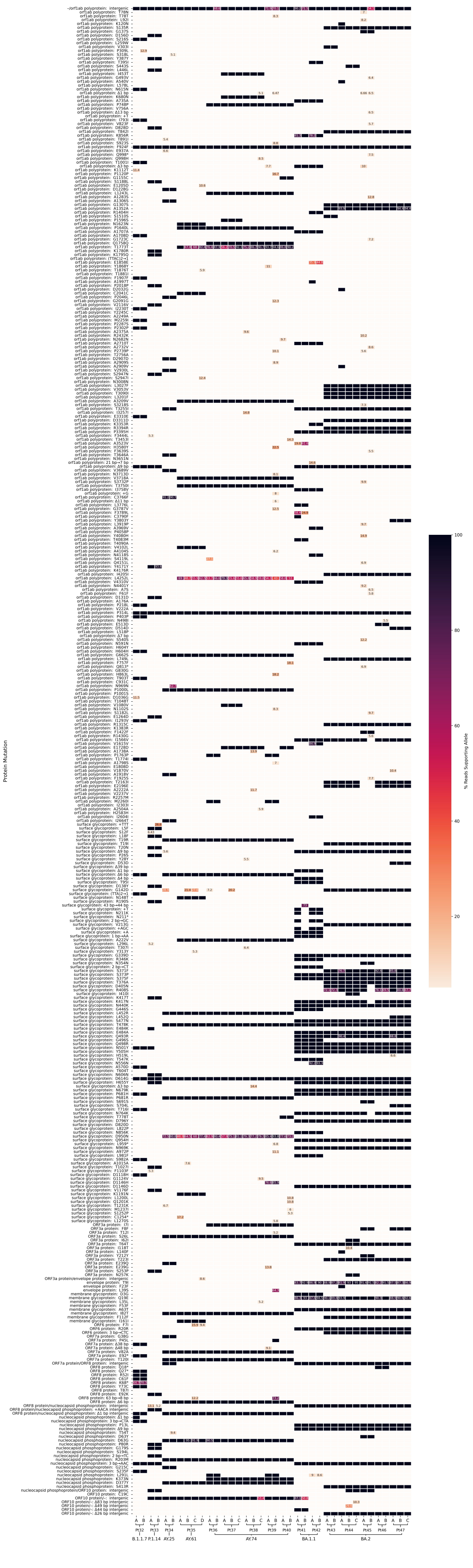

Protein Mutation
% Reads Supporting Allele
100
80
60
40
20
B.1.1.7
P.1.14
AY.25
AY.61
AY.74
BA.1.1
BA.2

B)

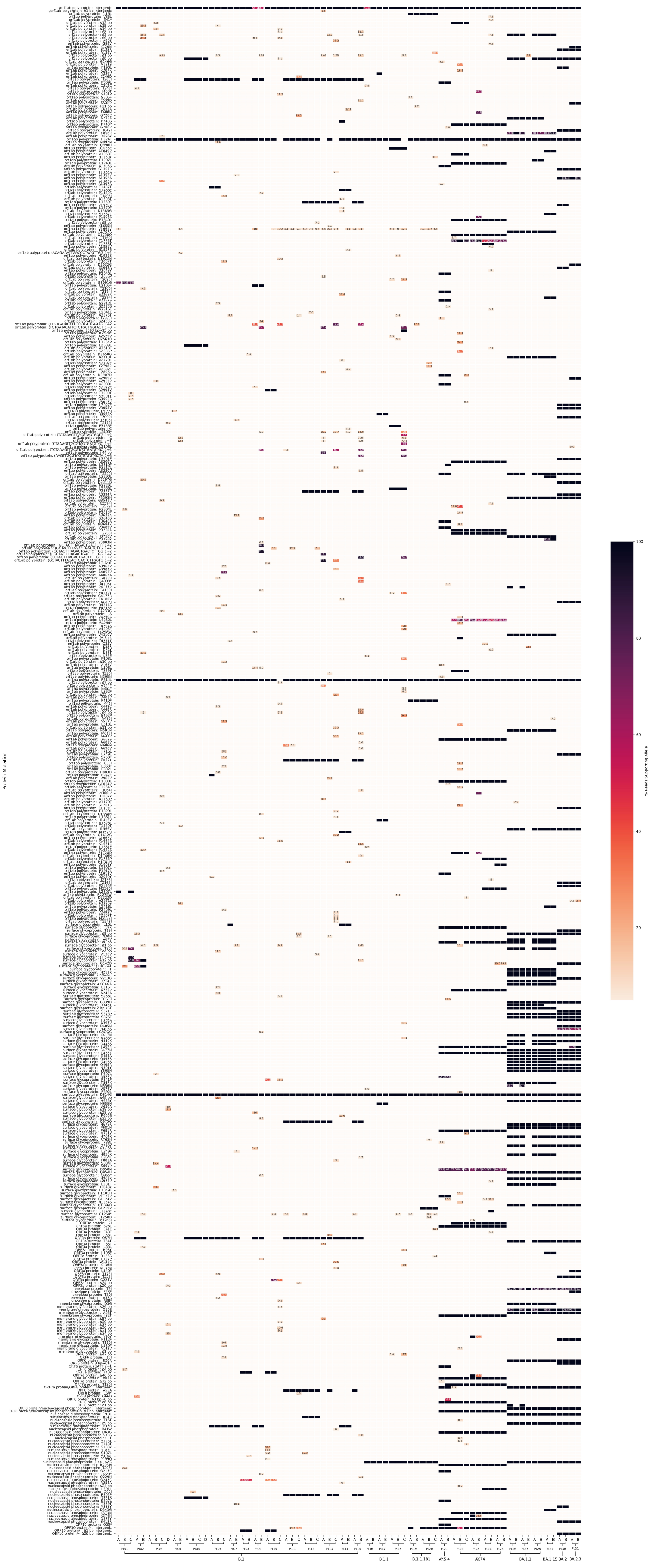
Protein Mutation
% Reads Supporting Allele
100
80
60
40
20
B.1
B.1.1
B.1.1.181
AY.5.4
AY.74
BA.1.1
BA.1.15
BA.2
BA.2.3

Supplementary Figure 1. Mutation profile across the whole genome for 39 serial swabs collected from RDV-treated patients (A) and 75 collected from untreated patients (B). Serial samples are labelled sequentially by date. Mutations were analyzed and called against the reference sequence with the Breseq pipeline. Low frequency minority mutations were retained and percentages are shown. Default parameters of Breseq including a 5% polymorphism frequency filter and a polymorphism minimum coverage of 2 were used.
